# U-Shaped Association Between Preoperative Fasting Blood Glucose and Postoperative Delirium in Elderly Diabetic Patients: A Multicentre Retrospective Cohort Study

**DOI:** 10.64898/2026.10.20.26366023

**Authors:** Xiaoying Zhang, Yun Zhang, Mengyao Yuan, Boyu Xu, Jiangbei Cao, Yanhong Liu, Jingsheng Lou, Wenzhu Shi, Qiang Fu, Weidong Mi

## Abstract

**Objective:** The heightened susceptibility of the aging brain to dysglycemia establishes blood glucose as a pivotal risk factor for postoperative delirium (POD) in elderly diabetic patients. This study investigated the association between preoperative fasting blood glucose (FBG) and POD risk in this population to define the optimal FBG range for POD.

**Methods:** This study included 2,157 elderly diabetic patients undergoing non-cardiac surgery between April 2020 and April 2022 in 20 hospitals across China. The POD incidence was prospectively assessed using the 3-minute Diagnostic Interview for Confusion Assessment Method (3D-CAM) within 7 postoperative days. The FBG-POD relationship was retrospectively analysed using restricted cubic splines within Cox proportional hazards model. Thresholds were validated with the machine learning-guided optimal cut-point analysis. A multivariable linear regression model was plotted to compare the distribution of time to POD onset and subgroup analyses were performed based on specific populations.

**Results:** A significant U-shaped relationship was observed between preoperative FBG and POD risk (*P* for nonlinearity=0.03). The hazard ratio identified the risk nadir of FBG at 8.43 mmol L^-1^. Optimal cut-point analysis defined three risk strata: a hypoglycemic-risk group (FBG ≤5.92 mmol L^-1^; 13.3% POD), an optimal-glycemic group (FBG 5.92-9.75 mmol L^-1^; 9.7% POD), and a hyperglycemic-risk group (FBG >9.75 mmol L^-1^; 18.5% POD). Compared with the optimal-glycemic group, both the hypoglycemic-risk group (HR=1.65, *P*<0.001) and the hyperglycemic-risk group (HR=1.51, *P*=0.03) showed significantly increased risk.

**Interpretation:** Preoperative FBG has a U-shaped association with POD risk in elderly diabetic patients with an optimal lowest-risk range as 5.92-9.75 mmol L^-1^.

**Summary Box:** *What is already known on this topic:*

- The aging brain is highly sensitive to dysglycemia, making elderly diabetic patients vulnerable to postoperative delirium.
- Current perioperative glucose management guidelines lack specific preoperative glycemic targets for preventing delirium in this population.

*What this study adds:*

- Preoperative fasting blood glucose (FBG) has a U-shaped association with postoperative delirium risk in elderly diabetic patients.
- An optimal neuroprotective FBG range of 5.92–9.75 mmol/L is associated with the lowest risk of delirium.
- Both hypoglycemia (≤5.92 mmol/L) and hyperglycemia (>9.75 mmol/L) independently increase delirium hazard by 65% and 51%, respectively.

## INTRODUCTION

The brain is characterized by the high demand for energy but low reserve capacity, a unique physiological profile that renders it exceptionally vulnerable to dysglycemia, potentially contributing to the occurrence of POD, especially in elderly Diabetes Mellitus (DM) patients. This sensitivity presents a double-edged sword: hypoglycemia directly induces energy crisis and excitotoxicity, whereas hyperglycemia triggers “glucotoxicity”, including osmotic stress, inflammatory cascades, oxidative stress, and vascular injury.^1,2^ The brain, as the primary target, suffers disruption of its intricate neural networks and neurotransmitter balance, culminating in acute, fluctuating disturbances of consciousness and cognitive deficits—manifesting as delirium. Consequently, elderly diabetic patients face a particularly challenging clinical dilemma during the perioperative period. However, the optimal preoperative glycemic target for preventing POD in elderly diabetic patients remains undefined.

Fasting blood glucose (FBG) is a low-cost, universally available, and routinely measured index of glycemic status. It serves as a practical and clinically actionable surrogate for assessing a patient’s metabolic stress level prior to undergoing surgical trauma.^3^ However, its relationship with POD, especially within the particularly vulnerable subgroup of elderly patients with diabetes, remains poorly defined and requires comprehensive investigation. Current perioperative blood glucose management guidelines lack a clear consensus on specific targets for elderly diabetic patients undergoing non-cardiac surgery. This study therefore aims to rigorously examine the association between preoperative FBG levels and the risk of POD in elderly diabetic patients, further to identify an optimal preoperative glycemic range associated with the lowest possible risk of delirium.

## METHODS

### Study Design and Population Selection

This investigation used a prospectively collected database and was conceived as a retrospective observational cohort study, conducted across 20 tertiary hospitals in China between April 10, 2020 and April 30, 2022.

The original prospective data collection was approved by the Ethics Committee of Chinese PLA General Hospital (Approval No. S2019-311-03). All patients enrolled in the original cohort provided written informed consent for the collection of their clinical data and for its use in future research. The present retrospective analysis of this prospectively collected database was separately approved by the Ethics Committee of Chinese PLA General Hospital (Approval No. S2025-699-01), and registered at Chinese Clinical Trial Registry (registration number: ChiCTR2500110735). Due to the retrospective, observational nature of this study, and because all data were de-identified prior to analysis, the requirement for additional informed consent for this specific analysis was waived by the ethics committee. The conduct and reporting of this study strictly adhered to the Strengthening the Reporting of Observational Studies in Epidemiology (STROBE) guidelines.^4^

The study population was derived from a cohort of 10,536 elderly patients who underwent non-cardiac surgery and had preoperative FBG data available. The inclusion criteria mandated that patients diagnosed with type 2 diabetes, be aged 65 yr or older, undergoing elective non-neurosurgical and non-cardiac procedures under general anaesthesia, and have at least two documented FBG measurements within the 30-day period preceding their surgery. To ensure a homogenous sample and minimize confounding, several exclusion criteria were applied. Exclusion criteria were: ASA physical status V; a history of major neurological disease (e.g., dementia, Alzheimer’s disease, Parkinson’s disease) or preoperative long-term use of antipsychotic medications; the mean of two most recent FBG value below 3.9 mmol L^-1^ or above13.9 mmol L^-1^; preoperative cognitive impairment (MMSE score <18); or missing data >5%.

### Exposure and Outcome Definitions

The primary exposure variable for this analysis was the preoperative fasting blood glucose level. This was defined as the mean of two most recent FBG value obtained from routine laboratory tests and recorded in the hospital’s electronic medical record system within the 30-day window before the index surgery.

The primary outcome was the incidence of postoperative delirium. POD in our prospectively collected database was diagnosed based on the criteria outlined in the Diagnostic and Statistical Manual of Mental Disorders, Fourth Edition (DSM-IV),^5^ occurring within the first seven days after surgery. Assessment of POD was conducted using the 3-Minute Diagnostic Interview for Confusion Assessment Method (3D-CAM),^6^ a widely recognized and validated instrument. Evaluations were performed face-to-face on the surgical wards by anaesthesia nurses who had received centralized, standardized training. Assessments were performed twice daily (at 10:00 AM and 5:00 PM) for 7 days postoperatively or until hospital discharge. The time of POD onset and duration was recorded as secondary outcomes of this current study. The duration of a delirium episode was defined as the number of days from its first recorded occurrence to its last. Other recorded clinical outcomes considered for analysis included cardiovascular complications, stroke, acute kidney injury, pneumonia, and the length of hospital stay.

### Statistical Analysis

The statistical analysis plan was comprehensive and multi-staged. Initially, patients were stratified into three preliminary groups based on clinically conventional FBG thresholds (≤7.7 mmol L^-1^, 7.8-10.0 mmol L^-1^, and >10.0 mmol L^-1^) to facilitate baseline characteristic comparisons.^7^ Categorical variables were presented as counts and percentages and compared using the Chi-square test or Fisher’s exact test, as appropriate. Continuous variables, which were non-normally distributed, were summarized as medians with interquartile ranges (IQRs) and compared using the Wilcoxon rank-sum test. Standardized mean differences (SMDs) were calculated to assess baseline balance across quintiles.

The core analysis investigating the relationship between FBG as a continuous variable and POD risk was performed using restricted cubic splines (RCS) with four knots, integrated within a Cox proportional hazards regression model. This non-parametric approach allows for the visualization of the relationship without assuming a linear form. The multivariable Cox model was adjusted for a pre-specified set of covariates selected based on clinical relevance and statistical considerations from univariate analyses. These covariates included patient age, sex, American Society of Anesthesiologists (ASA) physical status classification, significant preoperative weight loss (defined as >5 kg one month before surgery), the type of surgery performed, intraoperative administration of dexmedetomidine, and baseline cognitive and functional status as measured by the Mini-Mental State Examination (MMSE) and Basic Activities of Daily Living (BADL) scores. The proportional hazards assumption was verified using Schoenfeld residuals.

To quantitatively identify the inflection points of the observed nonlinear relationship, a two-piecewise Cox proportional hazards model was implemented. The threshold level was determined using an iterative process that maximized the model likelihood. Furthermore, an optimal data-driven cut-point analysis was employed to establish clinically actionable risk strata. This involved a machine learning-guided grid search algorithm that evaluated potential FBG thresholds between the 5th and 95th percentiles. For each candidate threshold, a Cox model was fitted, and the optimal cut-points were selected based on the Akaike Information Criterion (AIC)-adjusted Wald χ² statistic. The robustness and generalizability of these thresholds were subsequently validated using 10-fold cross-validation.

Subgroup analyses were pre-specified to examine the consistency and potential effect modification of the FBG-POD relationship. These analyses were stratified by age (≤75 yr vs. >75 yr), ASA classification (I-II vs. III-IV), receipt of perioperative blood transfusion (yes vs. no), surgical approach (open vs. endoscopic), and surgical specialty (e.g., thoracic, gastrointestinal, orthopedic). All statistical analyses were performed using R software (version 4.1.0), and a two-sided *P*-value of < 0.05 was considered statistically significant.

### Patient and public involvement

Patients or the public were not involved in the design, conduct, reporting, or dissemination plans of this research. This study is a retrospective analysis of a prospectively collected multi-centre cohort database, and the research question and outcome measures were developed based on clinical knowledge and scientific literature, rather than direct patient input. The results of this study will be disseminated through peer-reviewed publications and presentations at academic conferences.

## RESULTS

### Baseline Characteristics of the Study Cohort

We analysed all available data with a sample size of eligible 2,157 elderly patients with type 2 diabetes mellitus. Figure 1 illustrates the patient selection process. The incidence of POD was notably higher in this diabetic subpopulation (11.8%) compared to their non-diabetic counterparts (10.4%). The diabetic cohort had a median age of 72.1 yr (IQR: 68-77) and was predominantly male (53.8%). The majority of patients (67.5%) were classified as having ASA physical status I or II, indicating normal, healthy elderly patients and the ones with mild systemic disease.

**Figure 1.**
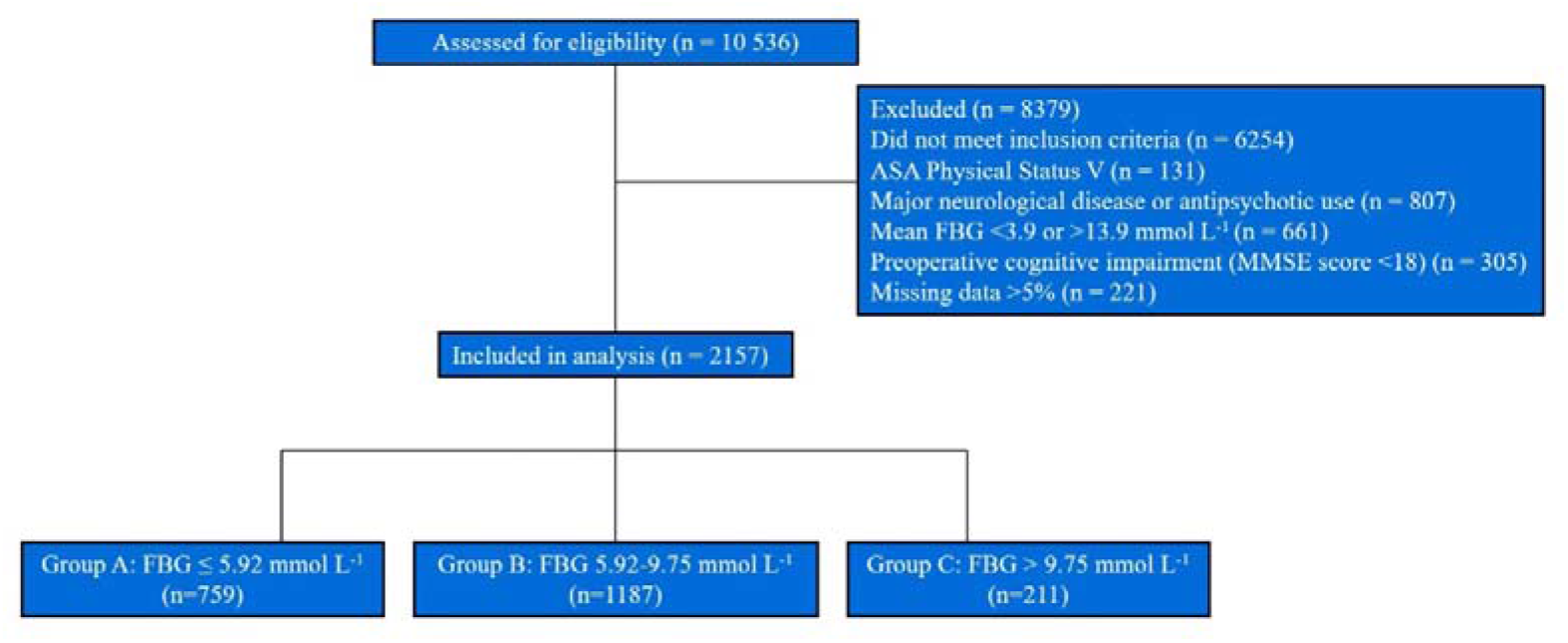
Flowchart of patient selection. Abbreviations: FBG, fasting blood glucose; ASA, American Society of Anesthesiologists; MMSE, Mini-Mental State Examination; POD, postoperative delirium.

A preliminary analysis stratifying patients by conventional FBG categories revealed a statistically significant difference in POD incidence (χ²=14.29, *P*<0.001). The incidence was 11.5% in the low FBG group (≤7.7 mmol L^-1^), 9.42% in the intermediate group (7.8-10.0 mmol L^-1^), and markedly higher at 19.6% in the high FBG group (>10.0 mmol L^-1^). Baseline comparisons among these groups showed no significant differences in terms of sex distribution, ASA classification, BMI category, educational level, or history of smoking and alcohol use (Supplementary Table S1).

### Nonlinear Relationship Between FBG and POD Risk

The Cox proportional hazards regression analysis first identified several independent predictors of POD (Supplementary Table S2). Advancing age was consistently associated with increased risk, with a 6% elevation in hazard per year in both univariate and multivariable models. Significant preoperative weight loss and specific surgical types, particularly hepatopancreatobiliary or gastrointestinal procedures, were also identified as significant risk factors. Conversely, intraoperative use of dexmedetomidine and higher baseline BADL scores were associated with a protective effect, reducing the hazard of delirium.

The RCS analysis was carried out and presented a statistically significant nonlinear relationship between preoperative FBG and the risk of POD (*P* for nonlinearity = 0.03). After full adjustment for the aforementioned covariates, the dose-response curve exhibited a distinct U-shape, indicating that both lower and higher extremes of FBG were associated with an elevated hazard of delirium. The proportional hazards assumption was satisfied for the overall model and for all individual covariates (Figure 2).

**Figure 2.**
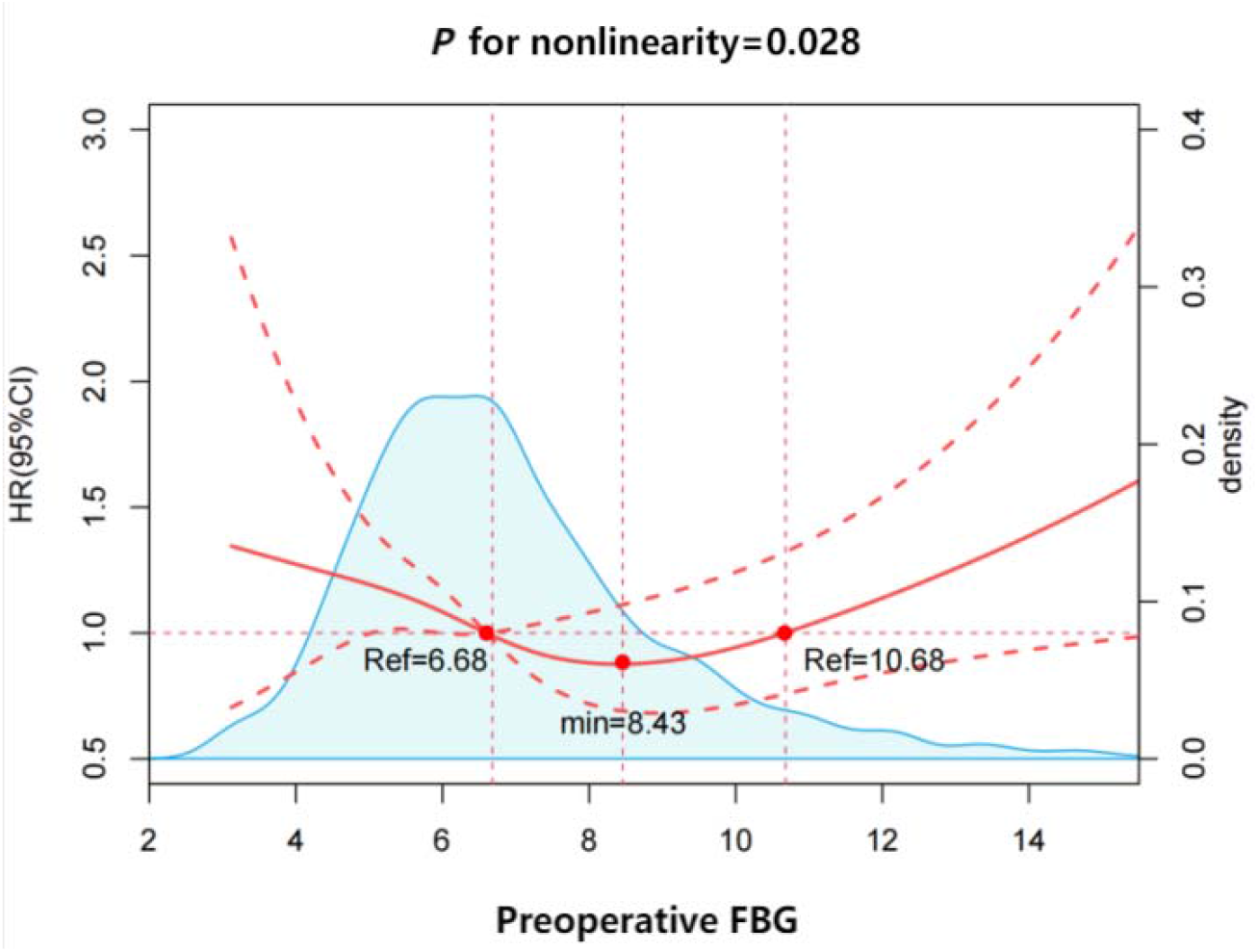
Restricted cubic spline curve showing the association between preoperative FBG and POD risk. Multivariable adjustment include age, sex, ASA classification, preoperative weight loss (>5 kg), type of surgery, intraoperative use of dexmedetomidine, preoperative MMSE score, and BADL score. A two-segment Cox proportional risk model was also used on both sides of the inflection point to explore the relationship between FBG and delirium risk. Abbreviations: FBG, fasting blood glucose; POD postoperative delirium; ASA, American Society of Anesthesiologists; MMSE, Mini-Mental State Examination; BADL, Basic Activities of Daily Living; HR, hazard ratio; CI, confidence interval.

Subsequent threshold effect modeling pinpointed an inflection point in this U-shaped curve at an FBG level of 8.43 mmol L^-1^. Below this threshold, each 1 mmol L^-1^ decrease in FBG was shown to be associated with a 13% increase in the hazard of POD (HR = 0.87, 95% CI: 0.78 to 0.98, *P=*0.02). In contrast, above this point of 8.43 mmol L^-1^, each 1 mmol L^-1^ increase in FBG was associated with a 10% increase in the hazard of POD (HR = 1.10, 95% CI: 1.00 to 1.20, *P=*0.04). Trend analyses across FBG quartiles further reinforced this bidirectional, non-monotonic relationship (Supplementary Table S3).

### Determination and Validation of Optimal Glycemic Thresholds

The machine learning-guided optimal cut-point analysis was used and identified two key FBG thresholds: 5.92 mmol L^-1^ and 9.75 mmol L^-1^. These values were used to stratify the cohort into three clinically relevant risk categories: Group A (Hypoglycemic Risk: FBG ≤ 5.92 mmol L^-1^ (n=759)) presented a POD incidence of 13.3%; Group B (Optimal Glycemic Range): FBG between 5.92 and 9.75 mmol L^-1^ (n=1,187)) demonstrated the lowest POD incidence of 9.7%; Group C (Hyperglycemic Risk: FBG > 9.75 mmol L^-1^ (n=211)) suffered the highest POD incidence of 18.5%. The differences in POD incidence among these three groups were highly significant (*P* < 0.001; Table 1). A comparative analysis of various candidate thresholds confirmed that the 5.92/9.75 mmol L^-1^ pair provided the best statistical discrimination for delirium risk (Supplementary Table S4).

**Table 1.**
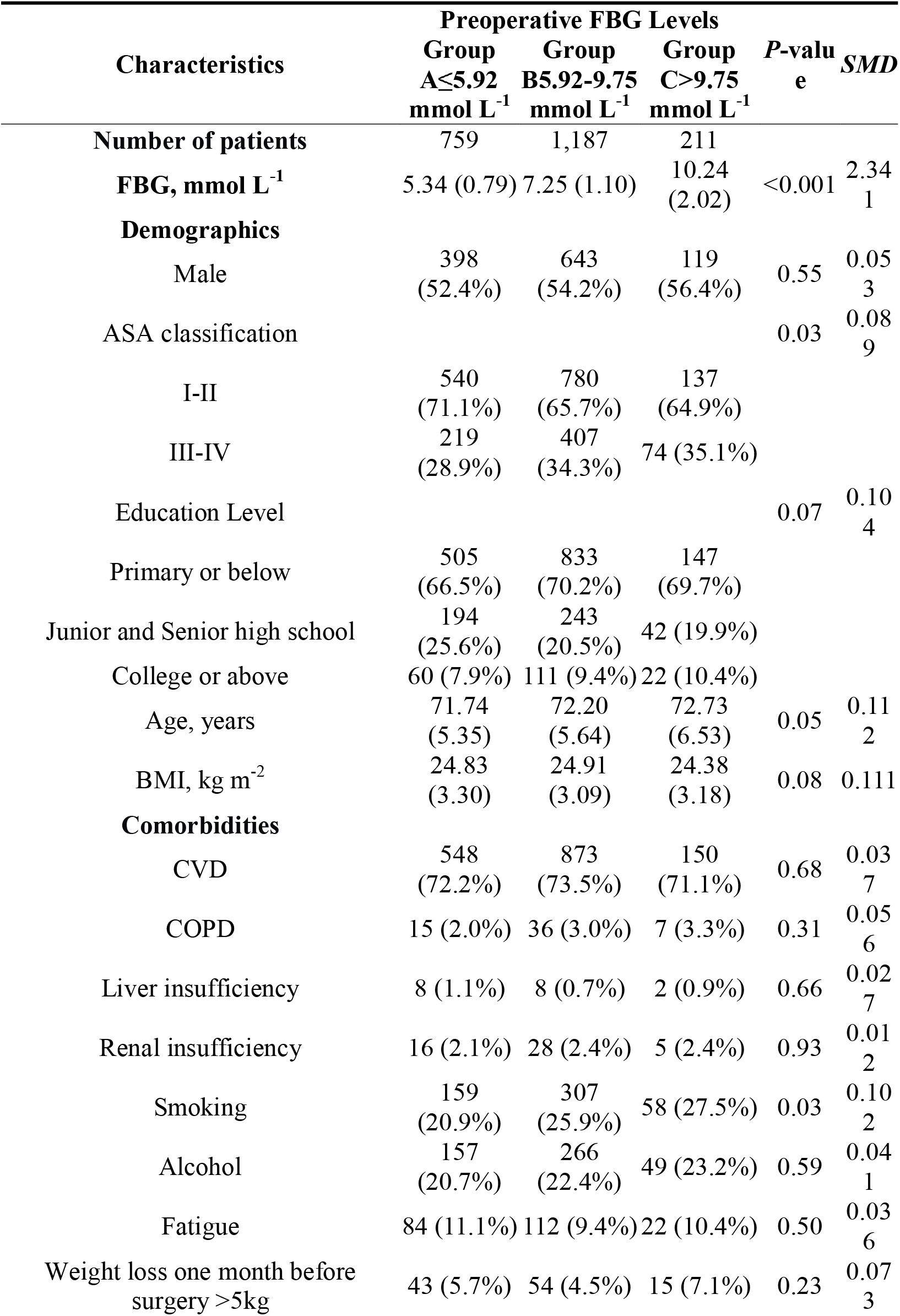
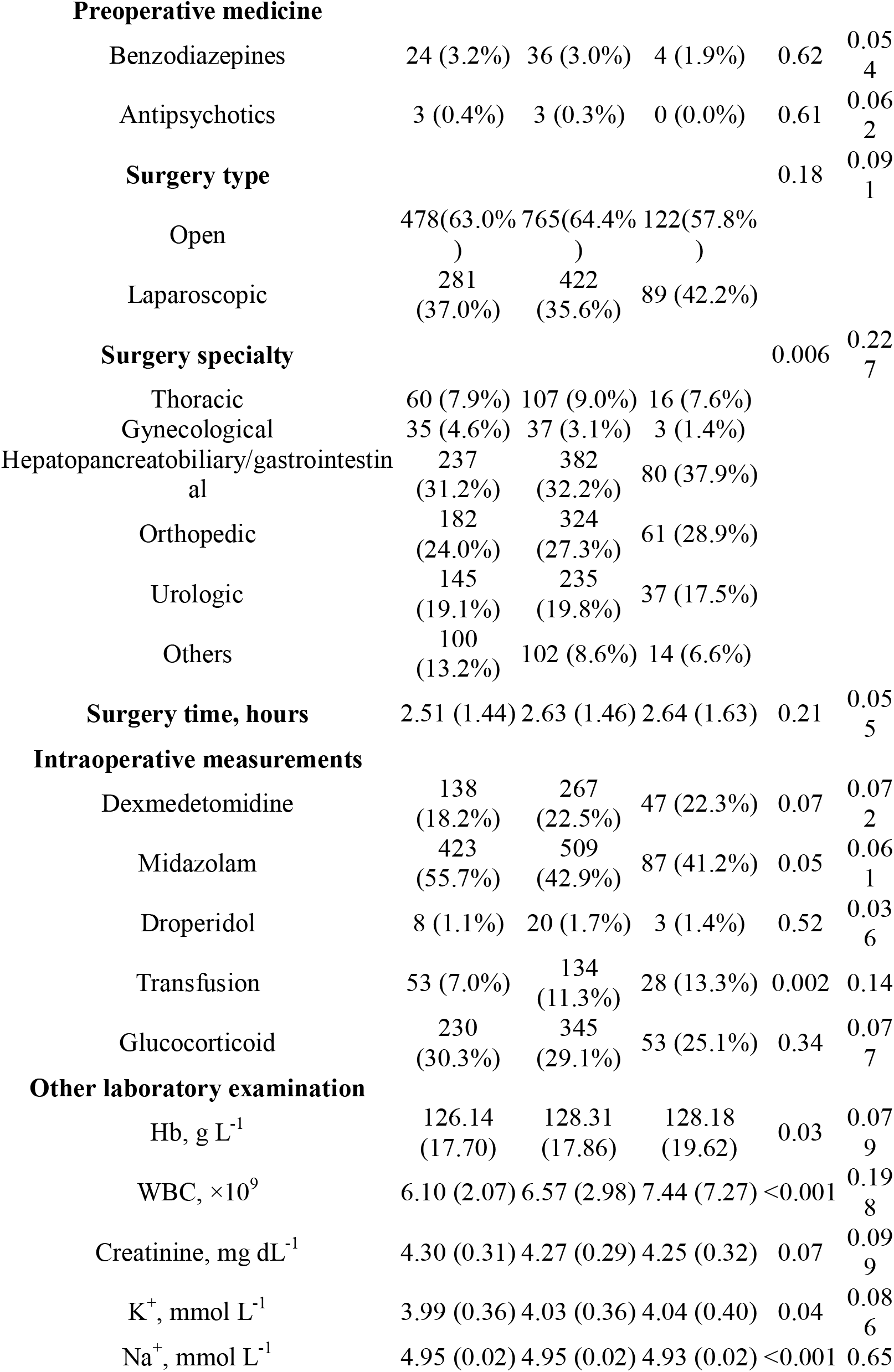
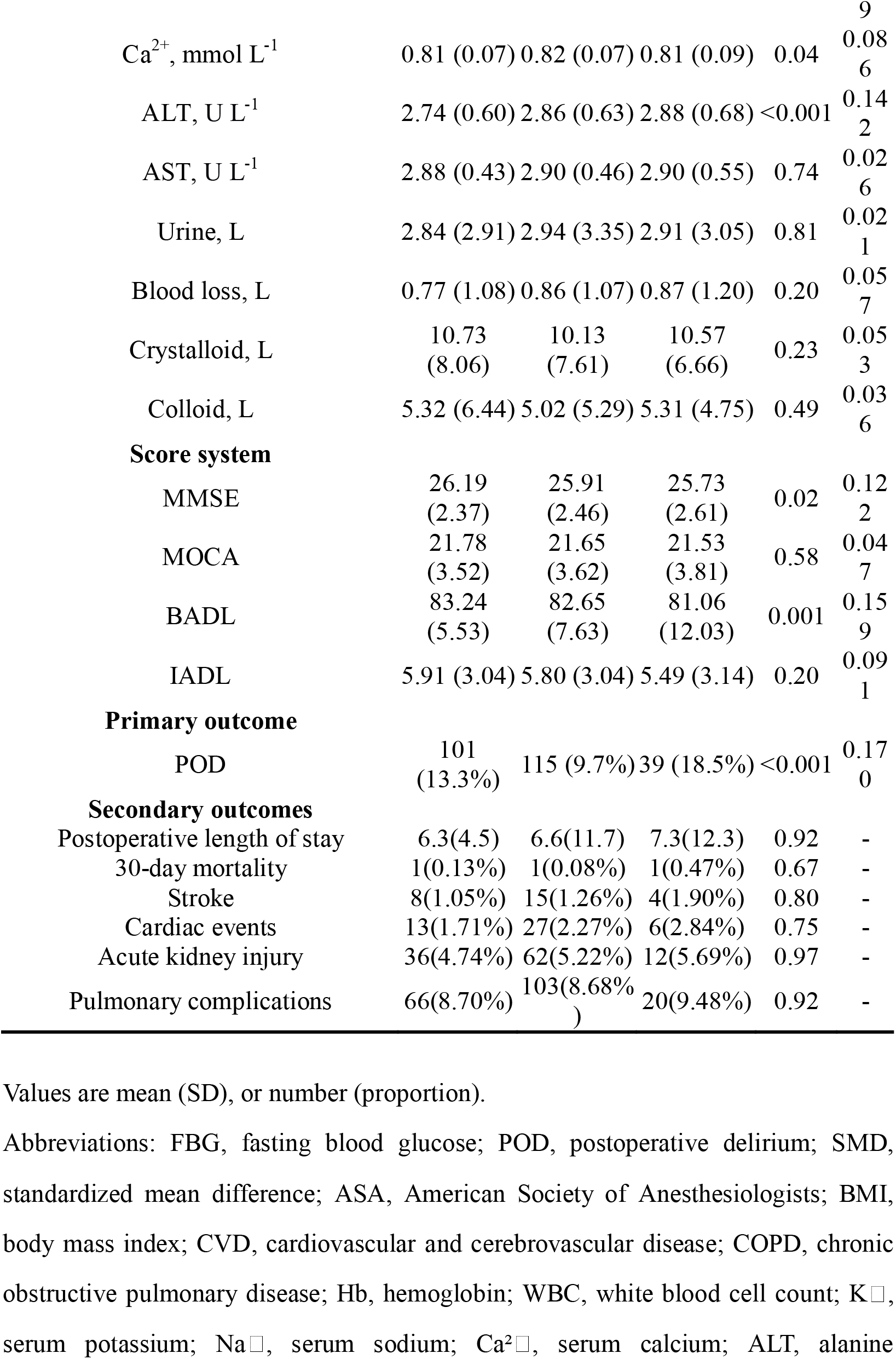
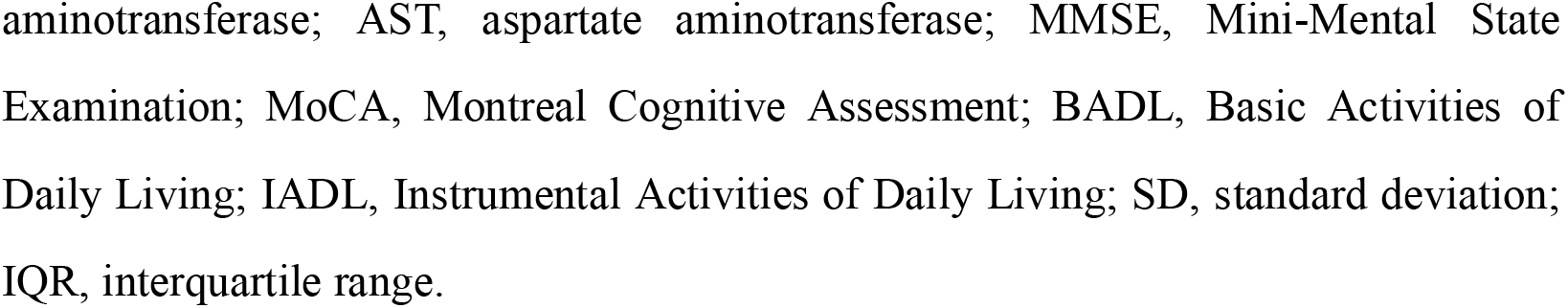
Characteristics and outcomes of participants categorized by threshold determination using the optimal cut-off points.

### Characteristics and Outcomes by Optimal Glycemic Strata

A detailed comparison of baseline characteristics across the three optimal FBG strata revealed intergroup differences. To quantitatively assess baseline balance, standardized mean differences (*SMD*s) were calculated and are reported in Table 1 alongside *P*-values. Most variables, including ASA physical status, age distribution, smoking history, types of surgery performed, rates of intraoperative transfusion, and various laboratory parameters (including hemoglobin, white cell count, and liver function indices), as well as baseline cognitive (MMSE) and functional (BADL) scores, showed minimal imbalance, suggesting that statistically significant *P*-values largely reflect the large sample size rather than clinically meaningful differences. However, to rigorously calibrate for intergroup confounding factors, these variables were incorporated as adjustment factors in the subsequent survival analysis.

Time-to-event analysis using Kaplan-Meier curves visually demonstrated a clear separation in delirium-free survival among the three groups over the 7-day postoperative period. Patients in Group B (optimal FBG) consistently exhibited the highest probability of remaining delirium-free. The log-rank test confirmed that these survival differences were statistically significant (*P* < 0.05) (Figure 3).

**Figure 3.**
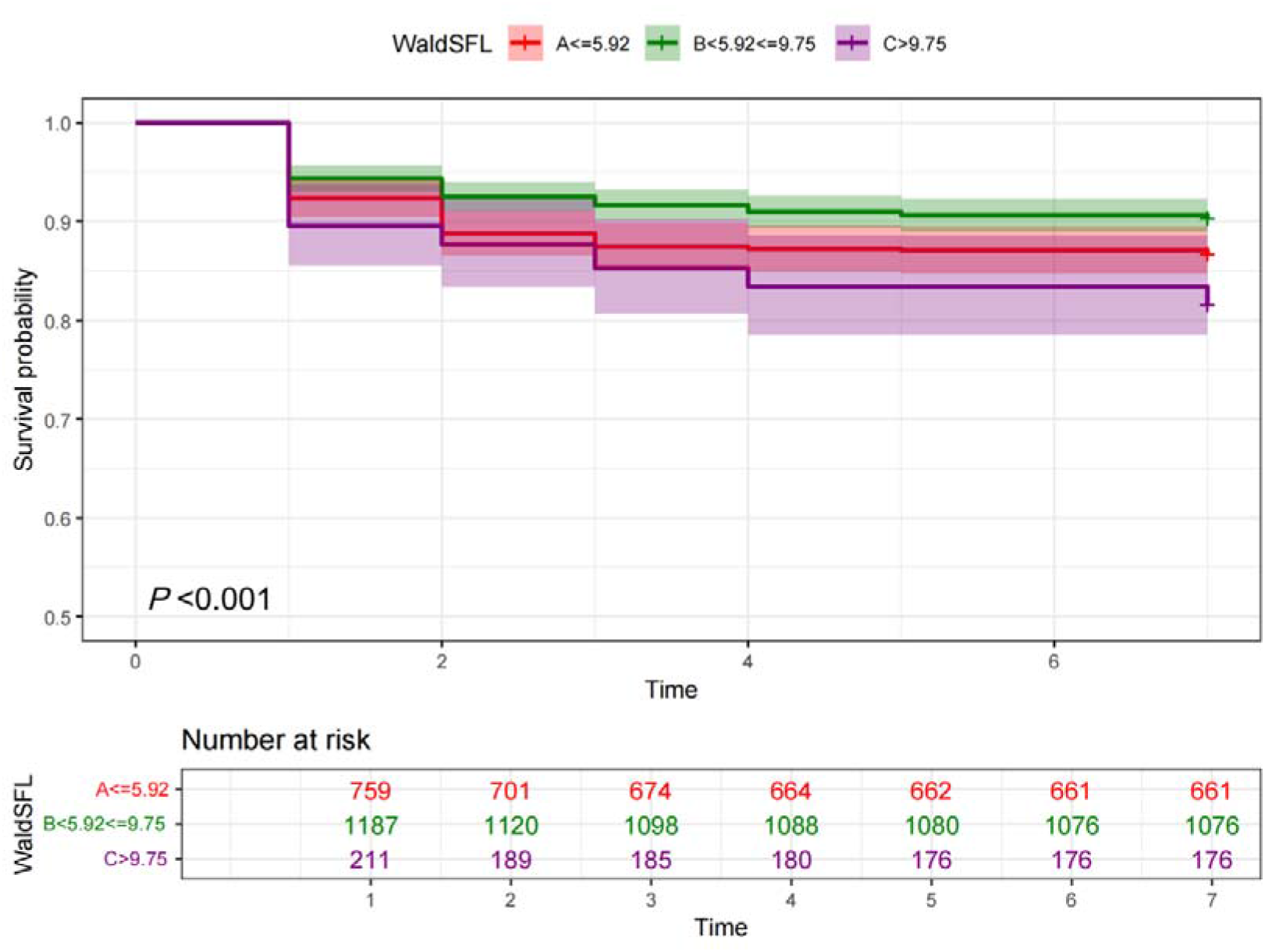
Kaplan-Meier survival analysis curves for POD in three groups of FBG.

In the final multivariate Cox proportional hazards regression model, after adjusting for all significant confounders including age, ASA status, surgical type, transfusion, MMSE, and BADL scores, both hypoglycemia and hyperglycemia remained independently associated with a substantially increased risk of POD. Using Group B as the reference, patients in Group A (FBG ≤ 5.92 mmol L^-1^) had a 65% higher hazard of delirium (HR = 1.65, 95% CI: 1.26 to 2.17, *P* < 0.001). Similarly, patients in Group C (FBG > 9.75 mmol L^-1^) had a 51% higher hazard (HR = 1.51, 95% CI: 1.03 to 2.21, *P* = 0.03) (Supplementary Table S5).

### Secondary Outcomes

After adjustment for potential confounding factors, there were no significant differences observed among the three glycemic strata for other secondary outcomes, including 30-day mortality, stroke, cardiac events, acute kidney injury, pulmonary complications, preoperative length of stay, duration of delirium, and time to delirium onset (all *P* > 0.05) (Table 1).

### Subgroup Analyses

The subgroup analyses provided deeper insights into the heterogeneity of the FBG-POD relationship. A pronounced, significant U-shaped curve (nonlinear *P* < 0.05) was consistently observed in clinically “robust” subgroups. These included patients aged between 65 and 75 yr, those with ASA I-II status, those who did not receive a perioperative blood transfusion, and those undergoing high-stress surgical procedures such as open surgery or specifically hepatopancreatobiliary/gastrointestinal operations. Conversely, in more “vulnerable” or physiologically compromised subgroups, the relationship transformed. For patients older than 75 yr, those with ASA III-IV status, those who received a blood transfusion, and those undergoing laparoscopic/luminal or orthopedic/gynecologic/urologic procedures, the U-shaped pattern was attenuated (Figure 4). In these subgroups, the relationship between FBG and POD risk often approximated a more linear, dose-response pattern, where the risk tended to increase steadily with rising FBG levels without a clear protective effect at the lower end of the spectrum.

**Figure 4.**
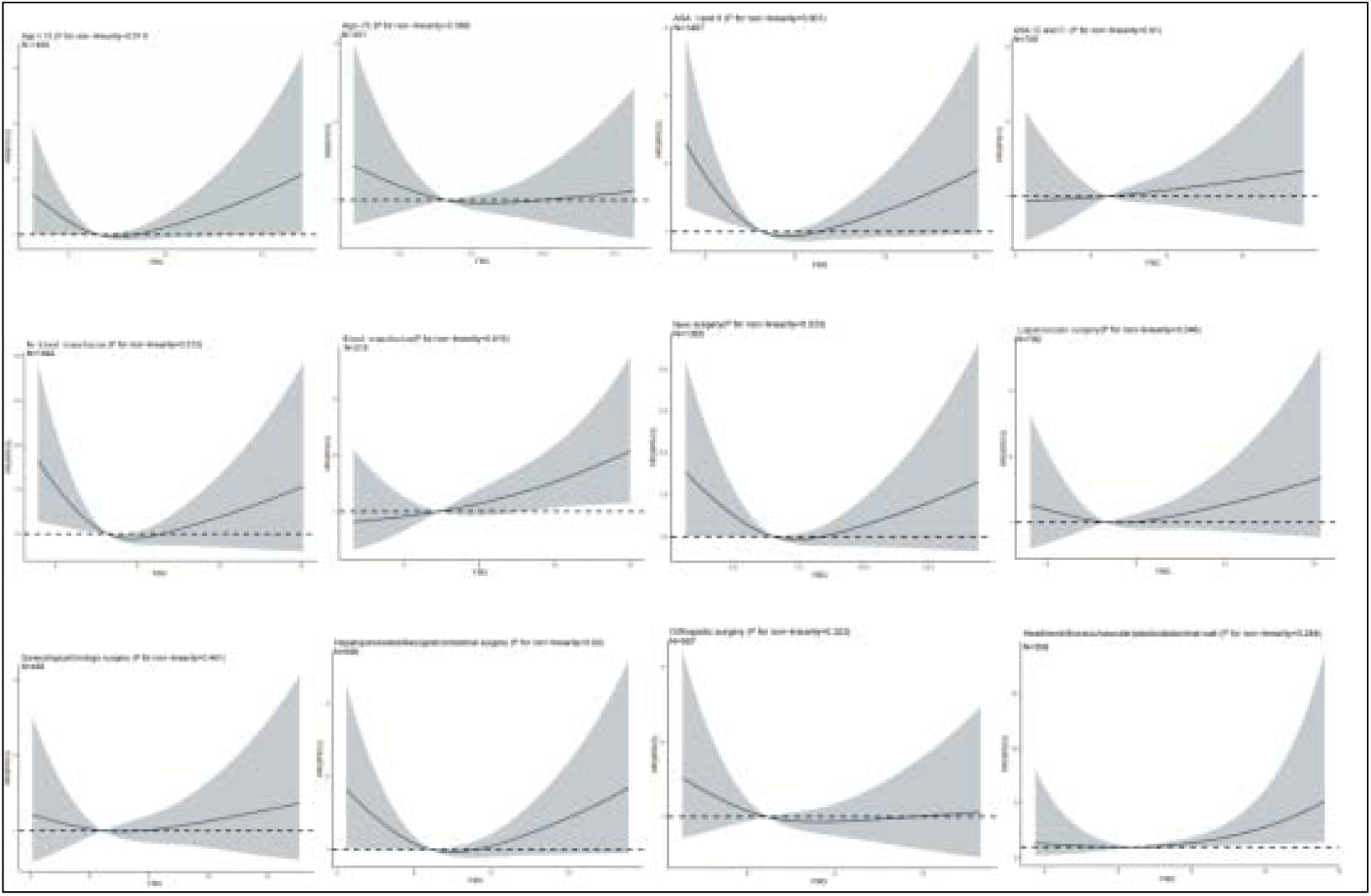
RCS curves for the relationship between FBG and POD in subgroup analyses. Multivariable adjustment include age, sex, ASA classification, preoperative weight loss (>5 kg), type of surgery, intraoperative use of dexmedetomidine, preoperative MMSE score, and BADL score. Abbreviations: FBG, fasting blood glucose; POD postoperative delirium; ASA, American Society of Anesthesiologists; MMSE, Mini-Mental State Examination; BADL, Basic Activities of Daily Living; HR, hazard ratio; CI, confidence interval.

## DISCUSSION

This multicenter cohort study utilized a prospectively collected database for retrospective analysis and revealed a significant, nonlinear, U-shaped association between preoperative fasting blood glucose levels and the risk of postoperative delirium in elderly patients with diabetes undergoing non-cardiac surgery. A distinct “neuroprotective” glycemic window is defined as an optimal preoperative FBG range between 5.92 and 9.75 mmol L^-1^.

Preoperative FBG, a widely available, low-cost, and routinely measured parameter, can be strategically repurposed as a valuable indicator for preoperative risk stratification and an easily achievable target for preventing postoperative delirium (POD). In patients identified preoperatively as falling outside the optimal range—specifically those with FBG ≤5.92 mmol L^-1^ or >9.75 mmol L^-1^—clinicians may consider implementing intensified perioperative monitoring, more frequent cognitive assessments, and proactive, meticulous glycemic management. This approach supports a shift in the goal of perioperative glucose management: from a reactive focus on avoiding extreme hyperglycemia or hypoglycemia toward a proactive strategy of precision neuroprotection, with the objective of maintaining blood glucose within a range optimized for brain health.

The biological plausibility of this U-shaped curve is supported by distinct yet converging pathophysiological pathways. On the hyperglycemic end, high glucose levels could facilitate the activation of microglia and release of the pro-inflammatory cytokines such as tumor necrosis factor-alpha (TNF-α) and interleukin-6 (IL-6).^8–10^ This triggers a state of central neuroinflammation, induces oxidative stress through the overproduction of reactive oxygen species (ROS), and disrupts the integrity of the blood-brain barrier.^11–13^ This cascade collectively damages synaptic plasticity and neuronal function, creating a fertile ground for delirium. On the hypoglycemic end, insufficient glucose delivery to the brain precipitates a cerebral energy crisis.^14^ Neurons, which are highly dependent on a continuous glucose supply, undergo depolarization, excitotoxicity, and ultimately, cellular injury. Brain regions critical for cognition, such as the hippocampus,^15^ are particularly vulnerable to these hypoglycemic insults,^16^ which are often compounded by pre-existing age-related or diabetic cerebral vulnerabilities. Furthermore, the impact of surgical stress, fasting, and energy deficit during prolonged surgery can aggravate severe glycemic variability.^17^

While our study identified a robust, U-shaped association between preoperative FBG and the risk of POD, the results did not show exactly same relationship in other postoperative complications with statistical significance. This dissociation underscores the specific neurotoxicity of dysglycemia in the vulnerable aging brain. The brain’s exceptional dependence on continuous glucose supply and its low threshold for metabolic and inflammatory injury may render it uniquely sensitive to preoperative glycemic extremes, even at levels that do not immediately precipitate systemic organ dysfunction. Of note, however, is that the group with the highest preoperative FBG (>9.75 mmol L ¹) still exhibited the highest incidence of all recorded complications, suggesting that perioperative FBG levels in elderly diabetic patients may still need to be maintained below 9.75 mmol L□¹.

Most intriguingly, the exploratory analysis of delirium kinetics suggested a divergent temporal pattern: an earlier onset trend with hypoglycemia and a later onset trend with hyperglycemia. This observation fuels the hypothesis that the two extremes of dysglycemia may precipitate delirium through distinct pathophysiological pathways with different time courses—an acute cerebral energy crisis versus a more protracted neuroinflammatory cascade.^18,19^ The lack of statistical significance for these secondary outcomes is likely multifactorial. First, our study was powered primarily for the primary outcome of POD, and the lower incidence of events like mortality or stroke limits statistical power to detect associations. Second, systemic complications are influenced by a vast array of intraoperative and postoperative factors (e.g., surgical technique, hemodynamic management, fluid balance) that may attenuate the isolated signal from a single preoperative metabolic snapshot. Third, our cohort consisted of elderly patients with diabetes, who inherently carry a high burden of vascular comorbidities; this background ‘noise’ may obscure the specific contribution of preoperative FBG to any single organ system’s outcome.

The subgroup analyses provide more in-depth evidence that paves the way for personalized perioperative medical care. The preservation of a clear U-shaped curve in patients with greater physiological reserve (e.g., younger age, better ASA status) suggests the existence of a metabolic “buffer zone” or “safety window”. Within this window, the body tend to tolerate and compensate for moderate glycemic fluctuations without tipping into delirium. However, deviations beyond this window likely trigger distinct pathological cascades (neuroinflammation or energy failure) that overwhelm compensatory mechanisms and precipitate the clinical syndrome of delirium. In contrast, for frailer patients (e.g., older, ASA III-IV, requiring transfusion), other significant perioperative factors diminish the impact of preoperative FBG on the occurrence of delirium, resulting in a flattened relationship in the RCS curve between preoperative FBG levels and delirium incidence.^20,21^

### Strengths and Limitations

The primary strengths of this study lie in its large, multicenter design to robustly model the nonlinear relationship and identify data-driven thresholds, enhancing the generalizability of the findings. The comprehensive adjustment for a wide array of potential perioperative confounders strengthens the validity of the observed associations.

However, the findings must be interpreted within the context of several limitations. The retrospective observational design is inherently susceptible to unmeasured confounding.^22^ Factors such as detailed diabetes duration, specific antidiabetic regimens, glycemic variability were not captured. Future efforts should focus on integrating data from preoperative fasting blood glucose, glycemic variability, and continuous glucose monitoring to build more precise risk prediction models to better guide the prevention and management of perioperative complications in elderly diabetic patients.

In conclusion, this study establishes a robust, U-shaped relationship between preoperative fasting blood glucose and the risk of postoperative delirium in elderly diabetic patients undergoing non-cardiac surgery. It delineates a neuroprotective glycemic window of 5.92 to 9.75 mmol L^-1^. Integrating this easily accessible indicator into preoperative assessment holds significant promise for enhancing risk stratification and guiding the development of individualized management plans.

## Declarations

### Ethics approval and consent to participate

This study was approved by the Ethics Committee of Chinese PLA General Hospital (Approval No. S2025-699-01) and registered at the Chinese Clinical Trial Registry (ChiCTR2500110735). All participants in the original cohort provided written informed consent. The requirement for additional informed consent for this specific retrospective analysis was waived by the ethics committee because all data were de-identified prior to analysis.

## Contributorship statement and guarantor

X-YZ, YZ, and M-YY contributed to data collection, methodology, investigation, and writing the original draft. W-ZS and J-SL contributed to methodology and investigation. Y-HL, J-BC, and B-YX contributed to data verification. Q-F contributed to conceptualization, project administration, supervision, and writing review and editing. W-DM contributed to conceptualization, project administration, supervision, funding acquisition, and writing review and editing. W-DM is the guarantor and accepts full responsibility for the work and the conduct of the study, had access to the data, and controlled the decision to publish. All authors approved the final version and meet the ICMJE authorship criteria.

## Data availability statement

The datasets generated and/or analysed during the current study are not publicly available due to restrictions imposed by the Ethics Committee of Chinese PLA General Hospital, as they contain information that could compromise participant privacy. De-identified data are available from the corresponding author on reasonable request, subject to approval from the institutional ethics committee.

## Transparency statement

The lead author (W-DM, the manuscript’s guarantor) affirms that the manuscript is an honest, accurate, and transparent account of the study being reported; that no important aspects of the study have been omitted; and that any discrepancies from the study as planned (and, if relevant, registered) have been explained.

## Dissemination plans

The results of this study will be disseminated through peer-reviewed publication and presentations at academic conferences. Patients and the public were not directly involved in the design of this study; therefore, no individual participant-level dissemination is planned.

