## supplementary material for "U-Shaped Association Between Preoperative Fasting Blood Glucose and Postoperative Delirium in Elderly Diabetic Patients: A Multicentre Retrospective Cohort Study"

**Table S1. Characteristics and outcomes of participants categorized by clinical preoperative FBG levels.**

| **Characteristics** | **Overall** | **Clinical Preoperative FBG Levels** | | | ***P*-value** |
| --- | --- | --- | --- | --- | --- |
|  |  | **≤7.7 mmol** **L^-1^** | **7.7–10 mmol** **L^-1^** | **>10mmol** **L^-1^** |  |
| **Number of patients** | 2,157 | 1,554 | 414 | 189 | 0.001 |
| **Demographics** |  |  |  |  |  |
| Male | 1160 (53.8%) | 825 (53.1%) | 229 (55.3%) | 106 (56.1%) | 0.58 |
| ASA classification |  |  |  |  | 0.29 |
| I-II | 1457 (67.5%) | 1065 (68.5%) | 269 (65.0%) | 123 (65.1%) |  |
| III-IV | 700 (32.5%) | 489 (31.5%) | 145 (35.0%) | 66 (34.9%) |  |
| Education Level |  |  |  |  |  |
| Primary or below | 1485 (68.8%) | 1061 (68.3%) | 291 (70.3%) | 133 (70.4%) | 0.54 |
| Junior and Senior high school | 479 (22.2%) | 354 (22.8%) | 90 (21.7%) | 35 (18.5%) | 0.54 |
| College or above | 193 (8.95%) | 139 (8.94%) | 33 (7.97%) | 21 (11.1%) | 0.54 |
| Age, years | 72.1 (5.64) | 71.9 (5.47) | 72.8 (5.87) | 72.4 (6.30) | 0.007 |
| BMI, kg m^-2^ | 24.8 (22.8,26.6) | 24.8 (22.8,26.6) | 24.8 (22.8,26.5) | 24.6 (22.4,26.4) | 0.1 |
| **Comorbidities** |  |  |  |  |  |
| CVD | 1571 (72.8%) | 1140 (73.4%) | 295 (71.3%) | 136 (72.0%) | 0.67 |
| COPD | 58 (2.69%) | 37 (2.38%) | 15 (3.62%) | 6 (3.17%) | 0.35 |
| Liver insufficiency | 18 (0.83%) | 13 (0.84%) | 3 (0.72%) | 2 (1.06%) | 0.85 |
| Renal insufficiency | 49 (2.27%) | 35 (2.25%) | 9 (2.17%) | 5 (2.65%) | 0.89 |
| Smoking | 524 (24.3%) | 360 (23.2%) | 112 (27.1%) | 52 (27.5%) | 0.15 |
| Alcohol | 472 (21.9%) | 331 (21.3%) | 95 (22.9%) | 46 (24.3%) | 0.54 |
| Fatigue | 218 (10.1%) | 157 (10.1%) | 43 (10.4%) | 18 (9.52%) | 0.95 |
| Weight loss( >5kg ) | 112 (5.19%) | 81 (5.21%) | 17 (4.11%) | 14 (7.41%) | 0.24 |
| **Preoperative medicine** |  |  |  |  |  |
| Benzodiazepines | 64 (2.97%) | 47 (3.02%) | 14 (3.38%) | 3 (1.59%) | 0.47 |
| Antipsychotics | 6 (0.28%) | 5 (0.32%) | 1 (0.24%) | 0 (0.00%) | 1 |
| **Surgery type** |  |  |  |  | 0.05 |
| Open | 1365 (63.3%) | 1007 (64.8%) | 250 (60.4%) | 108 (57.1%) |  |
| Laparoscopic | 792 (36.7%) | 547 (35.2%) | 164 (39.6%) | 81 (42.9%) |  |
| **Surgery specialty** |  |  |  |  | 0.05 |
| Thoracic | 183 (8.48%) | 132 (8.49%) | 38 (9.18%) | 13 (6.88%) |  |
| Gynecological | 75 (3.48%) | 63 (4.05%) | 9 (2.17%) | 3 (1.59%) |  |
| Hepatopancreatobiliary/gastrointestinal | 699 (32.4%) | 489 (31.5%) | 140 (33.8%) | 70 (37.0%) |  |
| Orthopedic | 567 (26.3%) | 391 (25.2%) | 120 (29.0%) | 56 (29.6%) |  |
| Urologic | 417 (19.3%) | 311 (20.0%) | 69 (16.7%) | 37 (19.6%) |  |
| Others | 216 (10.0%) | 168 (10.8%) | 38 (9.18%) | 10 (5.29%) |  |
| Surgery time, hours | 2.37 (1.58,3.28) | 2.38 (1.55,3.26) | 2.35 (1.60,3.40) | 2.33 (1.58,3.25) | 0.86 |
| **Intraoperative measurements** |  |  |  |  |  |
| Dexmedetomidine | 452 (21.0%) | 320 (20.6%) | 92 (22.2%) | 40 (21.2%) | 0.77 |
| Droperidol | 31 (1.44%) | 22 (1.42%) | 6 (1.45%) | 3 (1.59%) | 0.91 |
| Transfusion | 215 (9.97%) | 133 (8.56%) | 57 (13.8%) | 25 (13.2%) | 0.002 |
| Glucocorticoid | 628 (29.1%) | 456 (29.3%) | 125 (30.2%) | 47 (24.9%) | 0.38 |
| **Laboratory examination** |  |  |  |  |  |
| Hb, g L^-1^ | 128 (117,139) | 128 (117,139) | 130 (117,141) | 130 (115,141) | 0.31 |
| FBG, mmol L^-1^ | 6.84 (5.57,7.60) | 6.11 (5.30,6.87) | 8.36 (7.87,9.18) | 10.8 (9.73,11.9) | <0.001 |
| WBC, ×10^9^ | 6.21 (5.07,7.30) | 6.13 (5.00,7.10) | 6.36 (5.23,7.54) | 6.49 (5.25,8.11) | <0.001 |
| Creatinine, mg dL^-1^ | 4.28 (4.10,4.42) | 4.28 (4.11,4.43) | 4.27 (4.08,4.40) | 4.25 (4.04,4.44) | 0.07 |
| K^+^, mmol L^-1^ | 4.01 (3.78,4.22) | 4.01 (3.78,4.20) | 4.02 (3.77,4.27) | 4.02 (3.76,4.31) | 0.24 |
| Na^+^, mmol L^-1^ | 4.95 (4.94,4.96) | 4.95 (4.94,4.96) | 4.94 (4.93,4.95) | 4.94 (4.92,4.95) | <0.001 |
| Ca^2+^, mmol L^-1^ | 0.82 (0.79,0.85) | 0.82 (0.79,0.85) | 0.83 (0.80,0.86) | 0.82 (0.78,0.85) | 0.03 |
| ALT, U L^-1^ | 2.76 (2.42,3.09) | 2.75 (2.40,3.08) | 2.77 (2.48,3.18) | 2.78 (2.44,3.14) | 0.13 |
| AST, U L^-1^ | 2.84 (2.62,3.05) | 2.84 (2.63,3.05) | 2.83 (2.60,3.09) | 2.84 (2.54,3.13) | 0.62 |
| Urine, L | 2.13 (0.98,3.80) | 2.09 (0.96,3.73) | 2.28 (1.06,3.83) | 2.08 (0.98,4.17) | 0.27 |
| Blood loss, L | 0.46 (0.23,1.02) | 0.46 (0.23,1.00) | 0.51 (0.24,1.11) | 0.42 (0.18,1.10) | 0.18 |
| Crystalloid, L | 8.70 (6.35,12.1) | 8.59 (6.32,12.0) | 8.76 (6.41,12.2) | 8.93 (6.63,12.7) | 0.54 |
| Colloid, L | 3.78 (2.52,5.82) | 3.80 (2.55,5.87) | 3.68 (2.38,5.56) | 4.07 (2.56,6.28) | 0.33 |
| **Score system** |  |  |  |  |  |
| MMSE | 26 (26,27) | 26 (26,27) | 26 (26,27) | 26 (26,27) | 0.03 |
| MOCA | 21.7 (21.0,23.0) | 21.7 (21.0,24.0) | 21.7 (21.0,23.0) | 21.7 (21.0,23.0) | 0.54 |
| BADL | 85.0 (82.7,85.0) | 85.0 (82.7,85.0) | 85.0 (82.7,85.0) | 85.0 (82.7,85.0) | 0.18 |
| IADL | 7 (5,8) | 8(5,8) | 7 (4,8) | 7 (3,8) | 0.04 |
| **Outcome** |  |  |  |  | 0.001 |
| POD | 255 (11.8%) | 179 (11.5%) | 39 (9.42%) | 37 (19.6%) |  |

Values are mean(SD), number(proportion), or median(IQR).

Abbreviations: POD, postoperative delirium; ASA, American Society of Anesthesiologists; BMI, body mass index; CVD, cardiovascular and cerebrovascular disease; COPD, chronic obstructive pulmonary disease; FBG, fasting blood glucose; Hb, hemoglobin; WBC, white blood cell count; K⁺, serum potassium; Na⁺, serum sodium; Ca²⁺, serum calcium; ALT, alanine aminotransferase; AST, aspartate aminotransferase; MMSE, Mini-Mental State Examination; MoCA, Montreal Cognitive Assessment; BADL, Basic Activities of Daily Living; IADL, Instrumental Activities of Daily Living; SD, standard deviation; IQR, interquartile range.
**Table S2：Univariable and multivariable Cox proportional hazards model analysis of risk factors for POD.**

| **Characteristics** | **stats** | **Univariable**  **HR (95%CI)** | **Multivariable**  **HR (95%CI)** |
| --- | --- | --- | --- |
| Male | 1160 (53.8%) | 1.03 (0.80 to 1.32, *P*=0.83) | - |
| ASA classification |  |  |  |
| I-II | 1457 (67.5%) | 1 (Reference) | - |
| III-IV | 700 (32.5%) | 1.05 (0.81 to 1.36, *P*=0.71) | - |
| Education Level |  |  |  |
| Primary or below | 1485 (68.8%) | 1 (Reference) | - |
| Junior and Senior high school | 479 (22.2%) | 0.91 (0.67 to 1.24, *P*=0.56) | - |
| College or above | 193 (8.9%) | 1.26 (0.85 to 1.88, *P*=0.25) | - |
| Age, years | 72.1 (5.6) | 1.06 (1.04 to 1.08, *P*<0.001) | 1.06 (1.03 to 1.08, *P*<0.001) |
| BMI, kg m^-2^ | 24.8 (3.2) | 0.94 (0.90 to 0.98, *P*=0.002) | 0.97 (0.93 to 1.01, *P*=0.12) |
| CVD | 1571 (72.8%) | 1.23 (0.92 to 1.65, *P*=0.16) | - |
| COPD | 58 (2.7%) | 1.21 (0.60 to 2.44, *P*=0.60) | - |
| Liver insufficiency | 18 (0.8%) | 0.94 (0.23 to 3.77, *P*=0.93) | - |
| Renal insufficiency | 49 (2.3%) | 2.06 (1.12 to 3.76, *P*=0.02) | 1.55 (0.80 to 2.98, *P*=0.19) |
| Smoking | 524 (24.3%) | 1.03 (0.78 to 1.37, *P*=0.84) | - |
| Alcohol | 472 (21.9%) | 1.08 (0.81 to 1.44, *P*=0.61) | - |
| Fatigue | 218 (10.1%) | 1.33 (0.92 to 1.92, *P*=0.13) | - |
| Weight loss ( >5kg ) | 112 (5.2%) | 1.97 (1.29 to 3.00, *P*=0.002) | 1.51 (0.97 to 2.35, *P*=0.07) |
| Preoperative benzodiazepines | 64 (3.0%) | 1.21 (0.62 to 2.35, *P*=0.58) | - |
| Preoperative antipsychotics | 6 (0.3%) | 1.40 (0.20 to 10.01, *P*=0.74) | - |
| **Surgery type** |  |  |  |
| Open | 1365 (63.3%) | 1 (Reference) | - |
| Laparoscopic | 792 (36.7%) | 0.94 (0.72 to 1.21, *P*=0.62) | - |
| **Surgery specialty** |  |  |  |
| Thoracic | 183 (8.5%) | 1 (Reference) | 1 (Reference) |
| Gynecological | 75 (3.5%) | 0.22 (0.03 to 1.68, *P*=0.14) | 0.17 (0.02 to 1.34, *P*=0.09) |
| Hepatopancreatobiliary/gastrointestinal | 699 (32.4%) | 3.16 (1.71 to 5.86, *P*<0.001) | 2.16 (1.15 to 4.06, *P*=0.02) |
| Orthopedic | 567 (26.3%) | 1.77 (0.93 to 3.38, *P*=0.08) | 1.23 (0.63 to 2.38, *P*=0.54) |
| Urologic | 417 (19.3%) | 2.20 (1.15 to 4.21, *P*=0.02) | 1.88 (0.97 to 3.62, *P*=0.06) |
| Others | 216 (10.0%) | 0.45 (0.17 to 1.23, *P*=0.12) | 0.38 (0.14 to 1.03, *P*=0.06) |
| Surgery time, hours | 2.6 (1.5) | 1.21 (1.13 to 1.30, *P*<0.001) | 1.15 (1.05 to 1.25, *P*=0.001) |
| Intraoperative dexmedetomidine | 452 (21.0%) | 0.73 (0.52 to 1.01, *P*=0.06) | 0.65 (0.46 to 0.91, *P*=0.01) |
| Intraoperative midazolam | 1019(47.2%) | 0.98 (0.91 to 1.33, *P*=0.42) | - |
| Intraoperative droperidol | 31 (1.4%) | 1.10 (0.41 to 2.94, *P*=0.86) | - |
| Intraoperative transfusion | 215 (10.0%) | 1.25 (0.86 to 1.83, *P*=0.27) | - |
| Intraoperative glucocorticoid | 628 (29.1%) | 1.09 (0.84 to 1.43, *P*=0.51) | - |
| Hb, g L^-1^ | 127.5 (18.0 | 0.99 (0.98 to 0.99, *P*<0.001) | 1.00 (0.99 to 1.01, *P*=0.98) |
| FBG, mmol L^-1^ | 6.9 (1.8) | 1.04 (0.97 to 1.11, *P*=0.27) | - |
| WBC, ×10^9^ | 6.5 (3.4) | 1.01 (0.98 to 1.04, *P*=0.49) | - |
| Creatinine, mg dL^-1^ | 4.3 (0.3) | 1.50 (1.03 to 2.19, *P*=0.03) | 1.21 (0.83 to 1.76, *P*=0.33) |
| K^+^, mmol L^-1^ | 4.0 (0.4) | 1.02 (0.73 to 1.42, *P*=0.93) | - |
| Na^+^, mmol L^-1^ | 4.9 (0.0) | 0.00 (0.00 to 0.01, *P*<0.001) | 0.04 (0.00 to 14.82, *P*=0.29) |
| Ca^2+^, mmol L^-1^ | 0.8 (0.1) | 0.27 (0.07 to 0.95, *P*=0.04) | 0.22 (0.05 to 1.04, *P*=0.06) |
| ALT, U L^-1^ | 2.8 (0.6) | 1.21 (1.00 to 1.46, *P*=0.05) | 1.06 (0.78 to 1.43, *P*=0.70) |
| AST, U L^-1^ | 2.9 (0.5) | 1.49 (1.18 to 1.87, *P*<0.001) | 1.19 (0.81 to 1.75, *P*=0.37) |
| MMSE | 26.0 (2.4) | 0.93 (0.89 to 0.97, *P*=0.001) | 0.96 (0.91 to 1.01, *P*=0.09) |
| MOCA | 21.7 (3.6) | 0.94 (0.91 to 0.96, *P*<0.001) | 0.96 (0.93 to 0.99, *P*=0.004) |
| BADL | 82.7 (7.6) | 0.98 (0.97 to 0.99, *P*=0.002) | 0.99 (0.98 to 1.00, *P*=0.04) |
| IADL | 5.8 (3.0) | 0.95 (0.92 to 0.99, *P*=0.01) | 0.97 (0.93 to 1.01, *P*=0.12) |
| Urine, L | 2.9 (3.2) | 0.99 (0.95 to 1.03, *P*=0.62) | - |
| Blood loss, L | 0.8 (1.1) | 1.06 (0.96 to 1.16, *P*=0.28) | - |
| Crystalloid, L | 10.4 (7.7) | 0.99 (0.97 to 1.01, *P*=0.30) | - |
| Colloid, L | 5.2 (5.7) | 0.97 (0.94 to 1.00, *P*=0.05) | 1.00 (0.97 to 1.03, *P*=0.97) |
| FBG, mmol L^-1^ | 7.0 (2.3) | 1.04 (0.99 to 1.09, *P*=0.15) | - |

Values are mean(SD), number(proportion), or median(IQR).

Abbreviations: POD, postoperative delirium; ASA, American Society of Anesthesiologists; BMI, body mass index; CVD, cardiovascular and cerebrovascular disease; COPD, chronic obstructive pulmonary disease; FBG, fasting blood glucose; Hb, hemoglobin; WBC, white blood cell count; K⁺, serum potassium; Na⁺, serum sodium; Ca²⁺, serum calcium; ALT, alanine aminotransferase; AST, aspartate aminotransferase; MMSE, Mini-Mental State Examination; MoCA, Montreal Cognitive Assessment; BADL, Basic Activities of Daily Living; IADL, Instrumental Activities of Daily Living; FBG, fasting blood glucose.

**Table S3. Trend test and multivariable associations of preoperative FBG levels and POD risk across stratified thresholds.**

| **Characteristics** | **Preoperative FBG Levels** | | | |
| --- | --- | --- | --- | --- |
|  | **<8.43 mmol L^-1^**  **(n=1,740)** | | **>8.43 mmol L^-1^**  **(n=417)** | |
|  | **stats** | **HR (95%CI)** | **stats** | **HR (95%CI)** |
| Age | 71.9 (5.5) | 1.07 (1.05 to 1.10, *P*<0.001) | 72.9 (6.2) | 1.05 (1.00 to 1.09, *P*=0.04) |
| Male | 916 (52.6%) | 0.82 (0.61 to 1.10, *P*=0.19) | 244 (58.5%) | 1.33 (0.75 to 2.37, *P*=0.33) |
| ASA classification |  |  |  |  |
| I-II | 1187 (68.2%) | 1 (Reference) | 270 (64.7%) | 1 (Reference) |
| III-IV | 553 (31.8%) | 0.79 (0.58 to 1.09, *P*=0.16) | 147 (35.3%) | 0.96 (0.55 to 1.67, *P*=0.89) |
| Weight loss (>5 kg) | 89 (5.1%) | 1.73 (1.05 to 2.84, *P*=0.03) | 23 (5.5%) | 1.41 (0.58 to 3.38, *P*=0.45) |
| Surgery specialty |  |  |  |  |
| Thoracic | 149 (8.6%) | 1 (Reference) | 34 (8.2%) | 1 (Reference) |
| Gynecological | 70 (4.0%) | 0.23 (0.03 to 1.96, *P*=0.18) | 5 (1.2%) | 0.00 (0.00 to Inf, *P*=0.99) |
| Hepatopancreatobiliary/gastrointestinal | 541 (31.1%) | 3.74 (1.63 to 8.57, *P*=0.002) | 158 (37.9%) | 1.46 (0.56 to 3.80, *P*=0.44) |
| Orthopedic | 451 (25.9%) | 1.85 (0.78 to 4.39, *P*=0.16) | 116 (27.8%) | 0.67 (0.23 to 2.01, *P*=0.48) |
| Urologic | 342 (19.7%) | 3.11 (1.31 to 7.37, *P*=0.01) | 75 (18.0%) | 0.79 (0.26 to 2.41, *P*=0.67) |
| Others | 187 (10.7%) | 0.37 (0.10 to 1.33, *P*=0.13) | 29 (7.0%) | 0.35 (0.07 to 1.91, *P*=0.23) |
| Intraoperative dexmedetomidine | 360 (20.7%) | 0.87 (0.60 to 1.26, *P*=0.47) | 92 (22.1%) | 0.23 (0.09 to 0.60, *P*=0.003) |
| Preoperative MMSE scores | 26.0 (2.4) | 0.94 (0.90 to 0.99, *P*=0.02) | 25.8 (2.4) | 0.94 (0.85 to 1.03, *P*=0.20) |
| Preoperative BADL scores | 83.1 (6.2) | 0.97 (0.96 to 0.99, *P*<0.001) | 81.2 (11.5) | 0.99 (0.97 to 1.01, *P*=0.27) |
| FBG | 6.1 (1.2) | 0.87 (0.78 to 0.98, *P*=0.02) | 10.6 (2.3) | 1.10 (1.00 to 1.20, *P*=0.04) |
| **Stratified thresholds trend test** | ***P* for trend = 0.033** | | ***P* for trend =0.040** | |

Values are mean(SD), or number(proportion). Multivariable adjustment include age, sex, ASA classification, preoperative weight loss (>5 kg) , type of surgery, intraoperative use of dexmedetomidine, preoperative MMSE score, and BADL score.

Abbreviations: POD, postoperative delirium; ASA, American Society of Anesthesiologists; MSE, Mini-Mental State Examination; BADL, Basic Activities of Daily Living; FBG, fasting blood glucose; HR, hazard ratio.

**Table S4. Threshold determination using the optimal cut-off points.**

|  | **Non-delirium** | **Delirium** | ***P*-value** |
| --- | --- | --- | --- |
| **Threshold 1 (5.92-9.75)** |  |  | <0.001 |
| Out range (N=968) | 828 (85.5%) | 140 (14.5%) |  |
| Inrange (N=1189) | 1074 (90.3%) | 115 (9.7%) |  |
| **Threshold 2 (5.92-10.1)** |  |  | <0.001 |
| Out range (N=937) | 801 (85.5%) | 136 (14.5%) |  |
| In range (N=1220) | 1101 (90.2%) | 119 (9.8%) |  |
| **Threshold 3 (6.24-9.75)** |  |  | 0.04 |
| Out range (N=1131) | 981 (86.7%) | 150 (13.3%) |  |
| In range (N=1026) | 921 (89.8%) | 105 (10.2%) |  |
| **Threshold 4 (6.24-10.1)** |  |  | 0.04 |
| Out range (N=1100) | 954 (86.7%) | 146 (13.3%) |  |
| In range (N=1057) | 948 (89.7%) | 109 (10.3%) |  |

**Table S5. Univariable and multivariable Cox proportional hazards model analysis for POD.**

| **Characteristics** | **Stats** | **Univariable**  **HR (95%CI)** | **Multivariable**  **HR (95%CI)** |
| --- | --- | --- | --- |
| FBG, mmol L^-1^ |  |  |  |
| 5.92-9.75 | 1187 (55.0%) | 1 (Reference) | 1 (Reference) |
| ≤5.92 | 759 (35.2%) | 1.40 (1.07 to 1.83, *P*=0.01) | 1.65 (1.26 to 2.17, *P*<0.001) |
| >9.75 | 211 (9.8%) | 1.99 (1.38 to 2.86, *P*<0.001) | 1.51 (1.03 to 2.21, *P*=0.03) |
| ASA classification |  |  |  |
| Ⅰ-Ⅱ | 1457 (67.5%) | 1 (Reference) | 1 (Reference) |
| Ⅲ-Ⅳ | 700 (32.5%) | 1.05 (0.81 to 1.36, *P*=0.71) | 0.83 (0.63 to 1.09, *P*=0.17) |
| Age, years | 72.1 (5.6) | 1.06 (1.04 to 1.08, *P*<0.001) | 1.06 (1.04 to 1.08, *P*<0.001) |
| Smoke | 524 (24.3%) | 1.03 (0.78 to 1.37, *P*=0.84) | 1.07 (0.79 to 1.43, *P*=0.67) |
| Surgery specialty |  |  |  |
| Thoracic | 183 (8.5%) | 1 (Reference) | 1 (Reference) |
| Gynecological | 75 (3.5%) | 0.22 (0.03 to 1.68, *P*=0.14) | 0.17 (0.02 to 1.32, *P*=0.09) |
| Hepatopancreatobiliary/gastrointestinal | 699 (32.4%) | 3.16 (1.71 to 5.86, *P*<0.001) | 2.48 (1.33 to 4.64, *P*=0.004) |
| Orthopedic | 567 (26.3%) | 1.77 (0.93 to 3.38, *P*=0.08) | 1.24 (0.64 to 2.41, *P*=0.52) |
| Urologic | 417 (19.3%) | 2.20 (1.15 to 4.21, *P*=0.02) | 1.87 (0.97 to 3.61, *P*=0.06) |
| Others | 216 (10.0%) | 0.45 (0.17 to 1.23, *P*=0.12) | 0.32 (0.12 to 0.88, *P*=0.03) |
| Intraoperative blood transfusion | 215 (10.0%) | 1.25 (0.86 to 1.83, *P*=0.24) | 0.97 (0.65 to 1.45, *P*=0.88) |
| Hb, g L^-1^ | 127.5 (18.0) | 0.99 (0.98 to 0.99, *P*<0.001) | 1.00 (0.99 to 1.00, *P*=0.42) |
| WBC, ×10^9^ | 6.5 (3.4) | 1.01 (0.98 to 1.04, *P*=0.49) | 1.00 (0.98 to 1.03, *P*=0.86) |
| K^+^, mmol L^-1^ | 4.0 (0.4) | 1.02 (0.73 to 1.42, *P*=0.93) | 1.06 (0.75 to 1.49, *P*=0.74) |
| Na^+^, mmol L^-1^ | 4.9 (0.0) | 0.00 (0.00 to 0.01, *P*<0.001) | 0.00 (0.00 to 1.88, *P*=0.08) |
| Ca^2+^, mmol L^-1^ | 0.8 (0.1) | 0.27 (0.07 to 0.95, *P*=0.04) | 0.32 (0.06 to 1.58, *P*=0.16) |
| ALT, U L^-1^ | 2.8 (0.6) | 1.21 (1.00 to 1.46, *P*=0.05) | 1.22 (1.02 to 1.45, *P*=0.03) |
| Preoperative MMSE scores | 26.0 (2.4) | 0.93 (0.89 to 0.97, *P*=0.001) | 0.95 (0.90 to 0.99, *P*=0.01) |
| Preoperative BADL scores | 82.7 (7.6) | 0.98 (0.97 to 0.99, *P*=0.002) | 0.98 (0.97 to 1.00, *P*=0.006) |

Values are mean(SD), or number(proportion). Multivariable adjustment include age, sex, ASA classification, preoperative weight loss (>5 kg), type of surgery, intraoperative use of dexmedetomidine, preoperative MMSE score, and BADL score.

Abbreviations: POD, postoperative delirium; FBG, fasting blood glucose; ASA, American Society of Anesthesiologists; Hb, hemoglobin; WBC, white blood cell count; K⁺, serum potassium; Na⁺, serum sodium; Ca²⁺, serum calcium; ALT, alanine aminotransferase; MMSE, Mini-Mental State Examination; BADL, Basic Activities of Daily Living; SD, standard deviation; IQR, interquartile range.

**Figure S1. Kaplan-Meier survival analysis curves for POD in three groups of FBG stratified by multivariable variables adjustment.**

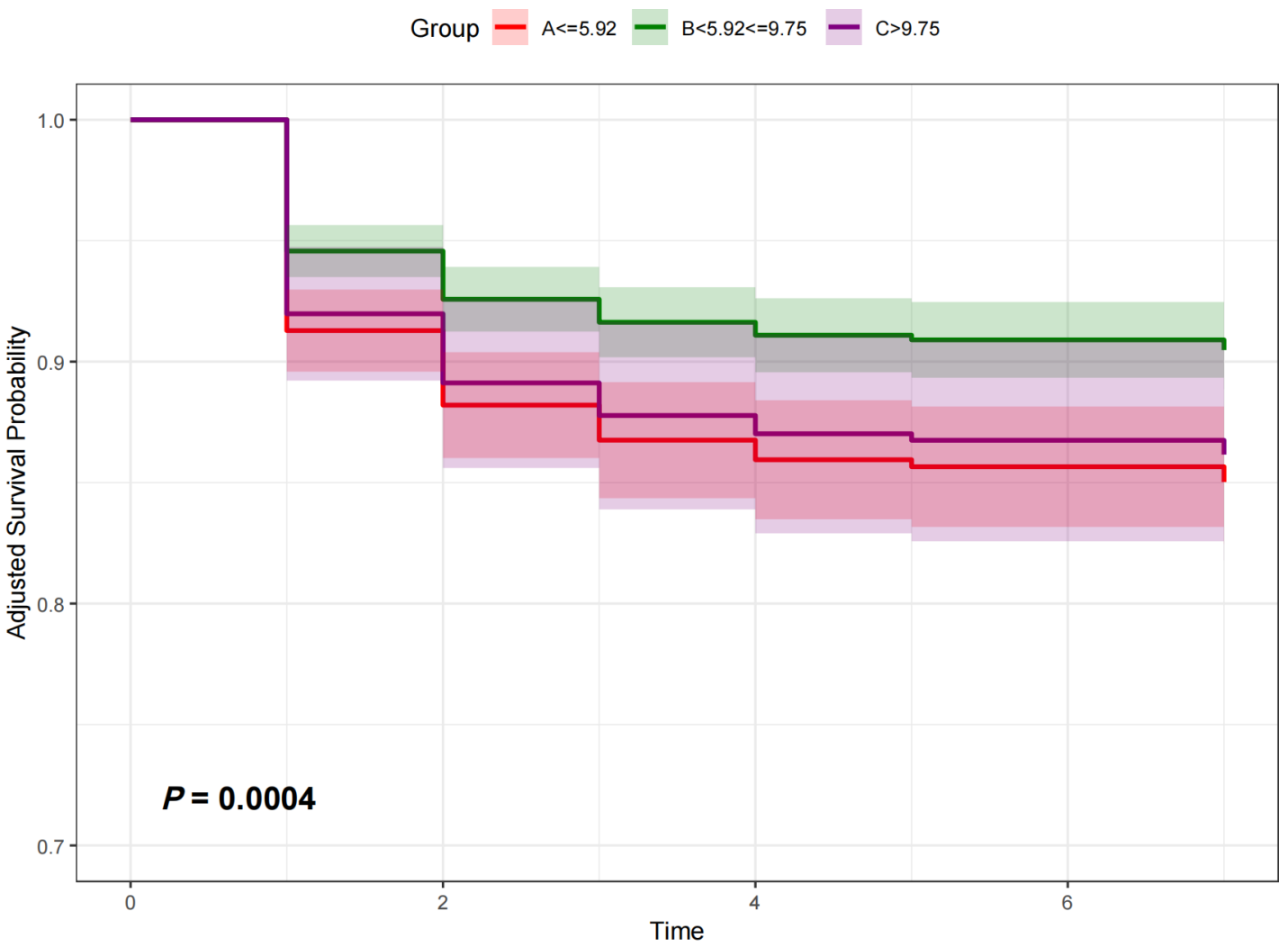

Multivariable adjustment include age, sex, ASA classification, preoperative weight loss (>5 kg), type of surgery, intraoperative use of dexmedetomidine, preoperative MMSE score, and BADL score. Abbreviations: POD postoperative delirium; FBG, fasting blood glucose; ASA, American society of anesthesiologists; MMSE, Mini-Mental State Examination; BADL, Basic Activities of Daily Living; HR, hazard ratio; CI, confidence interval**.**
